# Adjunctive GLP-1 Receptor Agonist Therapy Lowers Incident Pulmonary Hypertension and All-cause Mortality in Obese Patients with Obstructive Sleep Apnea Treated with PAP

**DOI:** 10.64898/2026.01.20.26344489

**Authors:** Samuel B. Governor, Ekow Essien, Abena K. Agyekum, Areeb Ahsan, Shimshon Wiesel, Dheeraj Khurana, Karim El-Kersh, Osamah Altaee, Prince Otchere, Victor G. Davila-Roman

## Abstract

**IMPORTANCE:** Obese subjects with obstructive sleep apnea (OSA) are at risk of pulmonary hypertension (PH) and mortality. Positive airway pressure (PAP) treatment lowers pulmonary artery pressures and mortality risk. Glucagon-like peptide-1 receptor agonists (GLP-1RAs) reduces body weight and cardiovascular risk, but their effectiveness in OSA remains unexplored.

**OBJECTIVE:** To assess whether GLP-1 RAs is associated with lower incident PH and all-cause mortality at 1 and 3 years in obese patients with OSA treated with PAP.

**DESIGN:** Retrospective cohort study.

**SETTING:** US Collaborative TriNetX Global Database analysis performed on January 5, 2026 encompassing data from 56 centers during December 1, 2017 and December 25, 2025.

**PARTICIPANTS:** Obese patients diagnosed with OSA and treated with PAP (n=288,587).

**EXPOSURE:** Propensity score-matched comparisons of PAP vs PAP+semaglutide, PAP vs PAP+tirzepatide, PAP+semaglutide vs PAP+tirzepatide.

**MAIN OUTCOMES AND MEASURES:** Incident PH and all-cause mortality at 1 and 3 years. Risk ratios (RRs) with 95% CIs were estimated using the delta method; number needed to treat (NNT) was calculated from the absolute risk difference.

**RESULTS:** Compared with PAP, PAP+semaglutide was associated with lower incident PH at 1 year (RR:0.45; 95% CI:0.44–0.56, NNT=38) and at 3years (RR:0.50; 95% CI:0.45–0.54, NNT=23), and lower mortality at 1 year (RR:0.35; 95% CI:0.33–0.37, NNT=12) and 3 years (RR:0.37; 95% CI:0.35–0.39, NNT=8); PAP+tirzepatide was associated with lower incident PH at 1 year (RR:0.27; 95% CI:0.22–0.34, NNT=28) and 3 years (RR:0.22; 95% CI:0.18–0.27, NNT=15) and lower mortality at 1 year (RR:0.14; 95% CI:0.12–0.17, NNT=9) and 3 years (RR:0.12; 95% CI:0.10–0.14, NNT=6). Compared to PAP+semaglutide, PAP+tirzepatide showed lower incident PH at 1 year (RR:0.51; 95% CI:0.40–0.65, NNT=77) and 3 years (RR:0.42; 95% CI:0.34–0.51, NNT=39) and lower all-cause mortality at 1 year (RR:0.41; 95% CI:0.34–0.49, NNT=40) and 3 years (RR:0.33; 95% CI:0.28–0.39, NNT=22); all p-values<0.001.

**CONCLUSIONS AND RELEVANCE:** Obese-OSA patients treated with PAP taking GLP-1RAs exhibited significantly lower 1-year and 3-year incident PH and all-cause mortality versus PAP. Tirzepatide exhibited further lowering of incident PH and all-cause mortality versus semaglutide, showing increased and sustained benefits over time.

**KEY POINTS:** *Question:* Among patients with obesity and obstructive sleep apnea (OSA) treated with positive airway pressure (PAP), is use of glucagon-like peptide-1 receptor agonists (GLP-1 RAs) associated with lower incident pulmonary hypertension (PH) and all-cause mortality at 1 and 3 years?

*Findings:* In a cohort study of 288,587 patients, PAP plus semaglutide or tirzepatide was associated with significantly lower incident PH and all-cause mortality versus PAP alone at 1 and 3 years. Tirzepatide was associated with lower risks than semaglutide.

*Meaning:* GLP-1 RAs may provide additional clinical benefit in obese patients with OSA treated with PAP.

## INTRODUCTION

Obstructive sleep apnea (OSA), a chronic disorder characterized by recurrent upper-airway collapse, hypoxemia, sympathetic activation, and cardiometabolic risk, affects more than 900 million people worldwide.^1,2^ Obesity is a central contributor to OSA, and weight loss strategies have resulted in improvements in apnea-hypopnea index (AHI) events and other disease biomarkers.^3,4^ Pulmonary hypertension (PH) is a major complication that develops in approximately 30-40% of OSA patients.^5^ Recurrent nocturnal hypoxia, sympathetic activation, negative intrathoracic pressure swings, oxidative stress, and systemic inflammation promote pulmonary vasoconstriction and vascular remodeling resulting in PH, while obesity contributes to PH by increased circulating volume, endothelial dysfunction, and increased left ventricular filling pressures.^6,7^ Consequently, in the setting of PH, individuals with obesity-related OSA exhibit increased morbidity (i.e., reduced exercise capacity, higher hospitalization risks) and mortality.^8,9^

Treatment of OSA with positive airway pressure (PAP) reduces hypoxemia, improves AHI, reduces pulmonary pressures and is associated with significantly lower all-cause mortality (37% reduction in risk versus untreated patients).^10,11^ Despite these salutary effects, the impact of PAP on PH seems heterogeneous and may be insufficient in patients with advanced metabolic disease or persistent hypoxic burden.^10,12^ Glucagon-like peptide-1 receptor agonists (GLP-1RAs) have emerged as highly effective agents for achieving clinically meaningful, sustained weight reduction, with additional benefits on blood pressure, inflammation, endothelial function and glycemic control. Randomized trials, including the SURMOUNT-OSA program, have shown that GLP-1RAs significantly reduce AHI, hypoxic burden and systemic inflammatory markers beyond weight reduction alone.^13-23^ These improvements extend to patients already receiving PAP, suggesting that pharmacologic weight loss may augment or complement ventilatory therapy by targeting the root mechanical and metabolic drivers of upper-airway collapse. Mechanistic and observational data also suggests that GLP-1RAs may confer pulmonary vascular benefits, potentially mitigating pathways relevant to PH development and progression, although definitive evidence in PH populations remains limited.

Given the persistent morbidity and mortality observed despite the use of guideline-directed PAP therapy in obese patients with OSA and PH, understanding the effects of novel therapeutic strategies on respiratory and cardiovascular systems represent a knowledge gap. To address this gap, the present study leverages a large federated electronic health record network database (TriNetX) to conduct statistical analysis of real-world data assessing the impact of GLP-1RAs in this high-risk obese population with OSA treated with PAP. The primary outcome is incident PH and all-cause mortality at 1-year and 3-years, comparing PAP versus PAP plus a GLP-1RA (either semaglutide or tirzepatide), and PAP plus semaglutide versus PAP plus tirzepatide.

## METHODS

### Data Source

This retrospective cohort study used the federated TriNetX Research Network database research platform electronic medical records from 56 healthcare centers across the United States. The network includes diagnoses, procedures, medications, and laboratory values collected during routine clinical care. All exposures, covariates, and outcomes were ascertained using identical data sources and coding definitions across all study cohorts. The TriNetX database is comprised of de-identified data, which complies with the Health Insurance Portability and Accountability Act (HIPAA) Privacy Rule and is exempt from institutional review board approval. The study followed the Strengthening the Reporting of Observational Studies in Epidemiology (STROBE) reporting guidelines.

### Study Population and Design

The overall study population was comprised of 288,587 adult patients (≥18 years) with obesity who met all three of the following criteria: a) obesity (body mass index [BMI]≥30 kg/m2); b) OSA (ICD-10-CM codes G47.33 and G47.30); and c) on treatment with PAP (CPT code 94660) between December 1, 2017 and December 25, 2025; data analysis was conducted on January 5, 2026. Patients either received or did not receive a GLP-1RA prescription during the study period and had at least two documented instances meeting the query criteria. After excluding 38,585 patients who did not meet entry criteria, the analytic sample of 250,002 patients was used to create three cohorts (Figure 1): a) PAP cohort (n=219,710) patients treated with PAP therapy who were not prescribed GLP-1RAs (semaglutide or tirzepatide) during the study period; 2) PAP+semaglutide cohort (n=21,149), patients treated with PAP therapy who were prescribed semaglutide but not tirzepatide; 3) PAP+tirzepatide cohort (n=9,143), patients treated with PAP therapy who were prescribed tirzepatide but not semaglutide.

**Figure 1.**
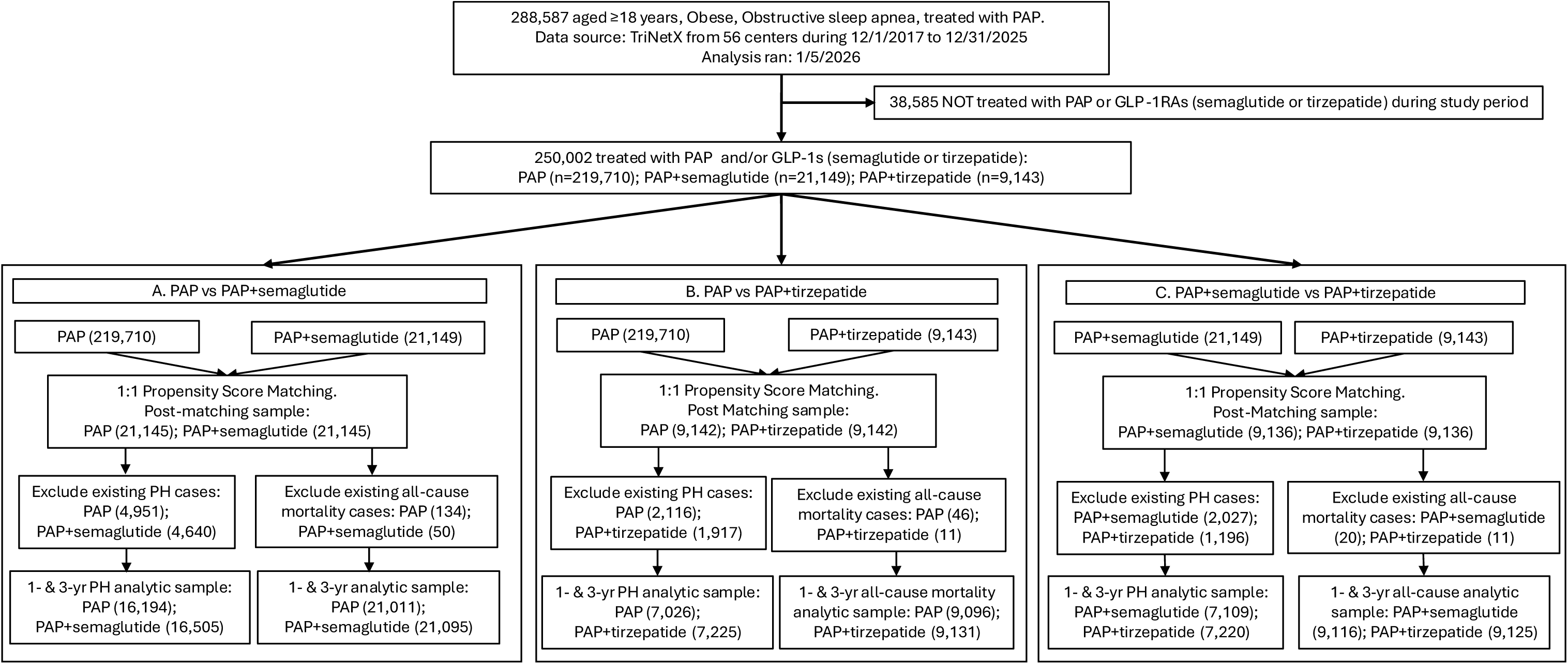
Flow Diagram of Cohort Construction.

For the 1- and 3-year analyses, patients with preexisting PH were excluded: A. PAP versus PAP+semaglutide (n=4,951 vs 4,640); B. PAP versus PAP+tirzepatide (n=2,116 vs 1,917); and C. PAP+semaglutide versus PAP+tirzepatide (n=2,027 vs 1,196). For all-cause mortality analyses, patients who died before the outcome assessment window were excluded: A. n=134 versus 50; B. n =46 versus 11; and C. n=20 versus 11, respectively.

The index date was defined as the date on which a patient first met the cohort eligibility and extended as an interval until all cohort criteria were satisfied. This approach resulted in a brief index period before initiation of the outcome assessment window. The outcome assessment window began 1 day after the index period ended and continued for 1 and 3 years, respectively for the 1-year and 3-year analyses. Patients were censored at the end of the prespecified follow-up period or at the time of outcome occurrence.

### Statistical Analysis

The study included 27 covariates encompassing demographics, comorbidities, and medications (full list of study variables in Table 1A and 1B). Continuous variables were assessed between comparison groups using independent-sample t-tests before and after matching, presented as means (sd); chi-square test was used to assess categorical variables between comparison groups before and after matching, presented as frequencies (percentages).

**Table 1A.**
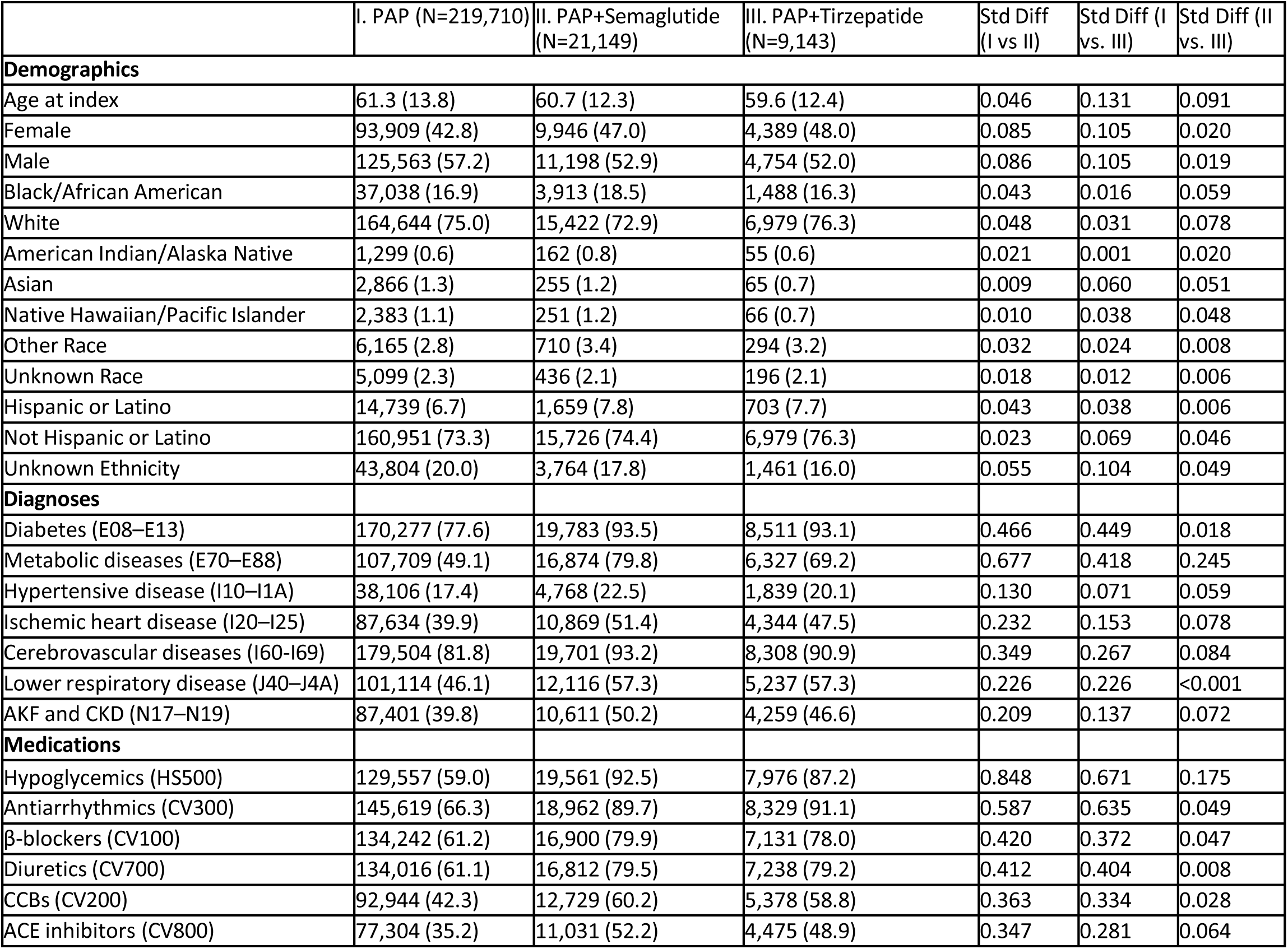
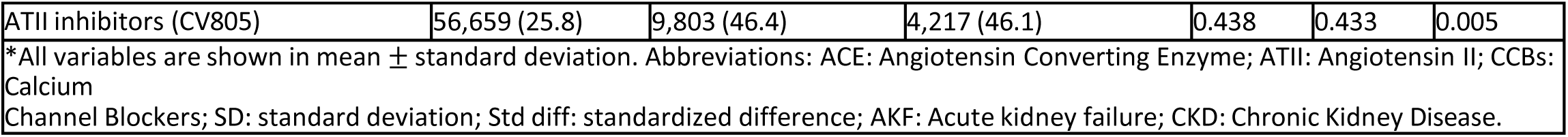
Baseline Characteristics for 1-year and 3-year Comparisons between PAP, PAP+Semaglutide and PAP+Tirzepatide Cohorts, Before Propensity Score Matching.

**Table 1B.** Baseline Characteristics for 1-year and 3-year Comparisons between PAP, PAP+Semaglutide and PAP+Tirzepatide Cohorts, After Propensity Score Matching.

|  |  |  |  |  |  |  |  |  |  |
| --- | --- | --- | --- | --- | --- | --- | --- | --- | --- |
|  | PAP vs. PAP+Semaglutide |  |  | PAP vs. PAP+Tirzepatide |  |  | PAP+Semaglutide vs. PAP+Tirzepatide |  |  |
|  | IV. PAP<br>(N=21,145) | V. PAP+Semaglutide<br>(N=21,145) | Std Diff<br>(IV vs V) | VI. PAP<br>(N=9,142) | VII. PAP+Tirzepatide<br>(N=9,142) | Std Diff<br>(VI vs VII) | VIII. PAP+Tirzepatide<br>(N=9,136) | IX. PAP+Semaglutide<br>(N=9,136) | Std Diff<br>(VIII vs IX) |
| **Demographics** |  |  |  |  |  |  |  |  |  |
| Age at index | 61.4 (12.6) | 60.7 (12.3) | 0.054 | 60.2 (13.2) | 59.6 (12.4) | 0.048 | 59.6 (12.4) | 59.8 (12.6) | 0.015 |
| Female | 9,765 (46.2) | 9,944 (47.0) | 0.017 | 4,255 (46.5) | 4,389 (48.0) | 0.029 | 4,386 (48.0) | 4,361 (47.7) | 0.005 |
| Male | 11,378 (53.8) | 11,198 (53.0) | 0.017 | 4,885 (53.4) | 4,753 (52.0) | 0.029 | 4,750 (52.0) | 4,772 (52.2) | 0.005 |
| Black/African American | 3,825 (18.1) | 3,913 (18.5) | 0.011 | 1,483 (16.2) | 1,488 (16.3) | 0.001 | 1,487 (16.3) | 1,464 (16.0) | 0.007 |
| White | 15,876 (75.1) | 15,418 (72.9) | 0.049 | 7,113 (77.8) | 6,978 (76.3) | 0.035 | 6,973 (76.3) | 7,038 (77.0) | 0.017 |
| Am Indian/Alaska Native | 114 (0.5) | 162 (0.8) | 0.028 | 45 (0.5) | 55 (0.6) | 0.015 | 55 (0.6) | 59 (0.6) | 0.006 |
| Asian | 220 (1.0) | 255 (1.2) | 0.016 | 47 (0.5) | 65 (0.7) | 0.025 | 65 (0.7) | 63 (0.7) | 0.003 |
| Nat Hawaiian/Pac Islander | 202 (1.0) | 251 (1.2) | 0.023 | 65 (0.7) | 66 (0.7) | 0.001 | 66 (0.7) | 64 (0.7) | 0.003 |
| Other Race | 573 (2.7) | 710 (3.4) | 0.038 | 247 (2.7) | 294 (3.2) | 0.030 | 294 (3.2) | 272 (3.0) | 0.014 |
| Unknown Race | 335 (1.6) | 436 (2.1) | 0.036 | 142 (1.6) | 196 (2.1) | 0.044 | 196 (2.1) | 176 (1.9) | 0.016 |
| Hispanic or Latino | 1,375 (6.5) | 1,658 (7.8) | 0.052 | 530 (5.8) | 703 (7.7) | 0.076 | 702 (7.7) | 670 (7.3) | 0.013 |
| Not Hispanic or Latino | 16,112 (76.2) | 15,723 (74.4) | 0.043 | 7,173 (78.5) | 6,978 (76.3) | 0.051 | 6,973 (76.3) | 7,056 (77.2) | 0.022 |
| Unknown Ethnicity | 3,658 (17.3) | 3,764 (17.8) | 0.013 | 1,439 (15.7) | 1,461 (16.0) | 0.007 | 1,461 (16.0) | 1,410 (15.4) | 0.015 |
| **Diagnoses** |  |  |  |  |  |  |  |  |  |
| Diabetes (E08–E13) | 19,839 (93.8) | 19,779 (93.5) | 0.012 | 8,579 (93.8) | 8,510 (93.1) | 0.031 | 8,504 (93.1) | 8,534 (93.4) | 0.013 |
| Metabolic dis (E70–E88) | 16,966 (80.2) | 16,870 (79.8) | 0.011 | 6,420 (70.2) | 6,326 (69.2) | 0.022 | 6,327 (69.3) | 6,335 (69.3) | 0.002 |
| Hypertensive disease (I10–I1A) | 4,740 (22.4) | 4,768 (22.5) | 0.003 | 1,834 (20.1) | 1,839 (20.1) | 0.001 | 1,839 (20.1) | 1,819 (19.9) | 0.005 |
| Ischemic heart disease (I20–I25) | 11,285 (53.4) | 10,869 (51.4) | 0.039 | 4,485 (49.1) | 4,344 (47.5) | 0.031 | 4,343 (47.5) | 4,460 (48.8) | 0.026 |
| Cerebrovascular disease | 19,776 (93.5) | 19,698 (93.2) | 0.015 | 8,346 (91.3) | 8,307 (90.9) | 0.015 | 8,303 (90.9) | 8,347 (91.4) | 0.017 |
| Lower respiratory disease (J40–J4A) | 12,061 (57.0) | 12,114 (57.3) | 0.005 | 5,201 (56.9) | 5,236 (57.3) | 0.008 | 5,231 (57.3) | 5,325 (58.3) | 0.021 |
| AKF and CKD (N17–N19) | 10,895 (51.5) | 10,611 (50.2) | 0.027 | 4,434 (48.5) | 4,259 (46.6) | 0.038 | 4,259 (46.6) | 4,409 (48.3) | 0.033 |
| **Medications** |  |  |  |  |  |  |  |  |  |
| Hypoglycemics (HS500) | 19,568 (92.5) | 19,557 (92.5) | 0.002 | 7,977 (87.3) | 7,975 (87.2) | 0.001 | 7,976 (87.3) | 8,019 (87.8) | 0.014 |
| Antiarrhythmics (CV300) | 19,105 (90.4) | 18,958 (89.7) | 0.023 | 8,377 (91.6) | 8,328 (91.1) | 0.019 | 8,322 (91.1) | 8,356 (91.5) | 0.013 |
| β-blockers (CV100) | 17,094 (80.8) | 16,897 (79.9) | 0.023 | 7,221 (79.0) | 7,130 (78.0) | 0.024 | 7,127 (78.0) | 7,235 (79.2) | 0.029 |
| Diuretics (CV700) | 17,078 (80.8) | 16,809 (79.5) | 0.032 | 7,356 (80.5) | 7,237 (79.2) | 0.032 | 7,233 (79.2) | 7,292 (79.8) | 0.016 |
| CCBs (CV200) | 13,025 (61.6) | 12,728 (60.2) | 0.029 | 5,464 (59.8) | 5,377 (58.8) | 0.019 | 5,373 (58.8) | 5,502 (60.2) | 0.029 |
| ACE inhibitors (CV800) | 11,195 (52.9) | 11,028 (52.2) | 0.016 | 4,633 (50.7) | 4,474 (48.9) | 0.035 | 4,474 (49.0) | 4,573 (50.1) | 0.022 |
| ATII Inhibitors (CV805) | 9,724 (46.0) | 9,801 (46.4) | 0.007 | 4,149 (45.4) | 4,216 (46.1) | 0.015 | 4,214 (46.1) | 4,270 (46.7) | 0.012 |
| *All variables are shown in mean ± standard deviation. Abbreviations: ACE: Angiotensin Converting Enzyme; ATII: Angiotensin II; CCBs: Calcium Channel Blockers; SD: standard deviation; Std diff: standardized difference; AKF: Acute kidney failure; CKD: Chronic Kidney Disease. | | | | | | | | | |

To minimize confounding by measured baseline characteristics and ensure comparability, 1:1 propensity score matching (PSM) was performed using a greedy nearest neighbor algorithm with a caliper width of 0.1 standard deviations for the following comparisons: PAP versus PAP+semaglutide, PAP versus PAP+tirzepatide, and PAP+semaglutide versus PAP+tirzepatide. PSM scores were estimated using multivariable logistic regression models that included baseline covariates, measured prior to or at the index date.

After PSM matching, covariate balance was assessed using standardized mean differences, with values <0.1 indicating adequate balance. All post-matching comparisons met this threshold, confirming successful balance of baseline characteristics between cohorts.

The two primary outcomes were incident pulmonary hypertension (PH), defined as a new diagnosis using ICD-10-CM codes I27.2, I27.20, I27.21, I27.22, I27.23, and I27.9, and all-cause mortality. Patients with outcomes occurring before the outcome assessment window were excluded using the TriNetX platform setting to exclude patients with prior outcomes. Thus, patients with preexisting PH and those who had died before the start of the outcome window were excluded.

Missing data were handled according to the TriNetX platform’s default complete-case analysis approach. No prespecified subgroup, interaction, or sensitivity analyses were performed. All analyses were conducted using R statistical software within the TriNetX platform. Outcomes were assessed during two prespecified time windows: 1 to 365 days after the index date (1-year analysis) and 1 to 1095 days after the index date (3-year analysis). Only incident cases were captured for outcome analyses. Six primary comparisons were performed: three for incident PH and three for all-cause mortality across the three cohorts at 1- and 3-years. For each outcome, the proportion of patients experiencing the event (risk) within each cohort was calculated. The number needed to treat (NNT) was calculated as the inverse of the absolute risk difference. Risk ratios (RRs) were calculated as the ratio of risks between cohorts, with 95% CIs estimated using the delta method. All statistical tests were two-sided, with p-values <0.05 considered statistically significant.

## RESULTS

### Study Population

A total of 288,587 adults with obesity and OSA treated with PAP were initially identified from the database. After application of prespecified cohort definitions and eligibility criteria, 250,002 patients were confirmed eligible and included in the analytic cohorts: a) PAP (n=219,710); b) PAP+semaglutide (n=21,149); and c) PAP+tirzepatide (n=9,143; Table 1A). After 1:1 propensity score matching, analytic cohorts included n= 21,145 for A) PAP versus PAP+semaglutide, n=9,142 for B) PAP versus PAP+semaglutide, n=9,136 for C) PAP+semaglutide versus PAP+tirzepatide. All matched cohorts demonstrated adequate covariate balance, with standardized mean differences <0.1 for all variables.

For analyses of incident PH, after excluding patients with a PH diagnosis prior to the start of the outcome window the final analytic samples were: 1) PAP (n=16,194) versus PAP+semaglutide (n=16,505); 2) PAP (n=7,026) versus PAP+tirzepatide (n=7,225); and 3) PAP+semaglutide (n=7,109) versus PAP+tirzepatide (n=7,220). For analyses of all-cause mortality, after excluding patients who died prior to the start of the outcome window the final analytic samples were: 4) PAP (n=21,011) versus PAP+semaglutide (n=21,095); 5) PAP (n=9,096) versus PAP+tirzepatide (n=9,131); and 6) PAP+semaglutide (n=9,116) versus PAP+tirzepatide (n=9,125). All outcomes were assessed over 1-year and 3-year follow-up periods.

Across all matched cohorts, the mean age was ∼60 years, with ∼52-53% being male and ∼70-73% White. The prevalence of comorbidities across cohorts was high, with cerebrovascular diseases and diabetes exceeding 90% prevalence and metabolic diseases exceeding 69% prevalence. Across cohorts, ∼48% or more had comorbid ischemic heart disease, lower respiratory disease, and acute kidney failure/chronic kidney disease, representing a very high-risk population (Table 1B).

### Primary outcomes

The primary outcomes consisted of analyses for incident PH at 1-year and 3-year follow-up and all-cause mortality at 1-year and 3-year follow-up (Table 2). Analyses of incident PH showed: 1) compared with PAP treatment, PAP+semaglutide treatment was associated with a markedly lower risk at both 1-year (RR:0.45; 95% CI:0.44–0.56, NNT=38) and 3-years (RR:0.50; 95% CI:0.45–0.54, NNT=23); 2) compared with PAP treatment, PAP+tirzepatide treatment was associated with a markedly lower risk of incident PH at both 1-year (RR:0.27; 95% CI:0.22–0.34, NNT=28) and 3-years (RR:0.22; 95% CI:0.18–0.27, NNT=15); 3) compared to PAP+semaglutide treatment, PAP+tirzepatide treatment was associated with a markedly lower risk at both 1-year (RR:0.51; 95% CI:0.40–0.65, NNT=77) and 3-years (RR:0.42; 95% CI:0.34–0.51, NNT=39). All estimates were derived from PSM matched cohorts and represent adjusted comparisons, and all analyses had p-value <0.001.

**Table 2.** Incident Pulmonary Hypertension and All-cause Mortality at 1-year and 3-years, by cohort.

|  |  |  |  |  |  |  |  |  |
| --- | --- | --- | --- | --- | --- | --- | --- | --- |
|  | Incident Pulmonary Hypertension |  |  |  | All-cause Mortality |  |  |  |
|  | No of events (%) | RR (95% CI),<br>p value,<br>NNT | No of events (%) | RR (95% CI),<br>p value,<br>NNT | No of events (%) | RR (95% CI),<br>p value,<br>NNT | No of events (%) | RR (95% CI),<br>p value,<br>NNT |
| Comparison I | 1 year |  | 3 year |  | 1 year |  | 3 year |  |
| PAP | 837/16,194 (5.2%) | 0.45 (0.44–0.56),<br>p < 0.001,<br>NNT = 38 | 1,425/16,194 (8.8%) | 0.50 (0.45–0.54),<br>p < 0.001,<br>NNT = 23 | 2,788/21,011 (13.3%) | 0.35 (0.33–0.37),<br>p < 0.001,<br>NNT = 12 | 4,212/21,011 (20.0%) | 0.37 (0.35–0.39),<br>p < 0.001,<br>NNT = 8 |
| PAP+Semaglutide | 425/16,505 (2.6%) |  | 720/16,505 (4.4%) |  | 975/21,095 (4.6%) |  | 1,549/21,095 (7.3%) |  |
| Comparison II | 1 year |  | 3 year |  | 1 year |  | 3 year |  |
| PAP | 348/7,026 (5.0%) | 0.27 (0.22–0.34),<br>p < 0.001,<br>NNT = 28 | 589/7,026 (8.4%) | 0.22 (0.18–0.27),<br>p < 0.001,<br>NNT = 15 | 1,138/9,096 (12.5%) | 0.14 (0.12–0.17),<br>p < 0.001,<br>NNT = 9 | 1,736/9,096 (19.1%) | 0.12 (0.10–0.14),<br>p < 0.001,<br>NNT = 6 |
| PAP+Tirzepatide | 96/7,225 (1.3%) |  | 134/7,225 (1.9%) |  | 162/9,131 (1.8%) |  | 205/9,131 (2.2%) |  |
| Comparison III | 1 year |  | 3 year |  | 1 year |  | 3 year |  |
| PAP+Semaglutide | 185/7,109 (2.6%) | 0.51 (0.40–0.65),<br>p < 0.001,<br>NNT = 77 | 318/7,109 (4.5%) | 0.42 (0.34–0.51),<br>p < 0.001,<br>NNT = 39 | 393/9,116 (4.3%) | 0.41 (0.34–0.49),<br>p < 0.001,<br>NNT = 40 | 622/9,116 (6.8%) | 0.33 (0.28–0.39),<br>p < 0.001,<br>NNT = 22 |
| PAP+Tirzepatide | 96/7,220 (1.3%) |  | 134/7,220 (1.9%) |  | 162/9,125 (1.8%) |  | 205/9,125 (2.2%) |  |
| Abbreviations: CI: confidence interval; PAP: positive airway pressure; NNT: number needed to treat. |  |  |  |  |  |  |  |  |

Analyses of all-cause mortality showed: 4) compared with PAP treatment, PAP+semaglutide treatment was associated with significantly lower risk at 1-year (RR:0.35; 95% CI:0.33–0.37, NNT=12) and 3-years (RR:0.37; 95% CI:0.35–0.39, NNT=8); 5) compared with PAP treatment, PAP+tirzepatide treatment showed significantly lower risk at 1-year (RR:0.14; 95% CI:0.12–0.17, NNT=9) and 3-years (RR:0.12; 95% CI:0.10–0.14, NNT=6); 6) Finally, PAP+tirzepatide was associated with a significantly lower mortality risk than PAP+semaglutide at both 1-year (RR:0.41; 95% CI:0.34–0.49, NNT=40) and 3-years (RR:0.33; 95% CI:0.28–0.39, NNT=22). All estimates were derived from propensity score-matched cohorts and represent adjusted comparisons, and all analyses had p-value <0.001.

## DISCUSSION

This study using a large federated database of obese patients with OSA who are treated with PAP at baseline in a real-world setting shows lower rates of incident pulmonary hypertension and all-cause mortality among those taking the GLP-1RAs semaglutide and tirzepatide. Furthermore, comparative effectiveness analysis between the two GLP-1RAs shows that those taking PAP+tirzepatide exhibited lower rates of incident PH and all-cause mortality compared to those taking PAP+semaglutide. The temporal trend analysis from 1-year to 3-years reveals sustained and increasing benefits, suggesting durable risk-reduction. These findings support the use of GLP-1RAs as adjunct treatment to PAP in obese patients with OSA, particularly those at very high cardiovascular risk such as the population studied. The differential efficacy between PAP+tirzepatide and PAP+semaglutide warrants further investigation into potentially distinct mechanistic pathways and may inform personalized therapeutic strategies in this high-risk population.

### OSA & PH - Epidemiology & Clinical Importance

Obesity and OSA are highly prevalent, incidence is increasing, and furthermore the prevalence of OSA rises steeply with increasing BMI.^3,4,24^ According to the American Academy of Sleep Medicine, “ten percent of weight gain is associated with a sixfold increase in risk of OSA”.^25^ Large population-and clinic-based studies show that markers of elevated pulmonary pressure by echocardiography are relatively common among people with moderate-to-severe OSA, particularly when nocturnal hypoxemia is frequent or prolonged.^6,26^ Risk is concentrated in patients with more severe AHI and greater nocturnal desaturation (i.e., greater hypoxic burden), older age, long duration of untreated OSA, coexisting chronic lung disease, and comorbid left-heart disease, all conditions that increase pulmonary venous pressure or chronically raise pulmonary vascular resistance.^7,26^

PH in the OSA population is heterogeneous in both mechanisms and prognosis. While the majority of patients have post-capillary or “combined” PH due to left-heart disease (i.e., from heart failure with preserved or reduced EF) and/or obesity-related left ventricular diastolic dysfunction, a smaller subset has predominantly hypoxia-driven or obstructive-sleep-related precapillary PH, while others have both. Regardless of the mechanism, the combination of OSA and PH is associated with worse functional status, higher healthcare utilization and poorer long-term outcomes compared with those who have OSA without PH.^27^ As PAP therapy reduces pulmonary pressures in patients with obesity and OSA, and is also associated with significantly lower all-cause mortality, the findings of the present study are potentially of high clinical impact as treatment with both PAP and GLP-1RAs was associated with lower incident PH and lower all-cause mortality.^10,11^

### GLP-1RA RCTs and Meta-analyses

In light of the high cardiometabolic risks in patients with concomitant obesity, OSA and PH, evaluation of treatment strategies addressing the mechanisms for both the respiratory disorder and its systemic vascular consequences represents a major clinical gap. Accordingly this study was designed to evaluate the potential benefits of GLP-1RAs on this population, as recent studies have shown value in this population. The strongest evidence to-date comes from the SCALE Sleep (liraglutide) and the more recent phase-3 SURMOUNT-OSA program (tirzepatide) RCTs in obese subjects with OSA.^15,28^. The SCALE Sleep RCT in participants with obesity and moderate/severe OSA showed that after 32 weeks, the liraglutide group exhibited a greater mean reduction in AHI than with placebo (-12.2 vs -6.1 events h-1, estimated treatment difference: -6.1 events h-1, 95% CI: -11.0 to - 1.2, P=0.015). Furthermore, liraglutide produced significantly greater reductions than placebo in body weight, systolic blood pressure (SBP) and glycated hemoglobin. The SURMOUNT-OSA study, comprised two 52-week, phase-3 RCTs in adults with obesity and moderate-to-severe OSA, including a sub-study of participants on stable PAP. In this study tirzepatide showed ∼16–17% mean weight loss and reduced AHI by ∼20–24 events/hour (∼48–56% relative reduction), decreased hypoxic burden and hsCRP, and improved SBP and sleep-related patient reported outcomes. Benefits were seen both in participants using PAP and those not using PAP.^23^

Evidence that GLP-1RAs reduce risk and/or severity of pulmonary hypertension is suggestive but currently is limited mostly by preclinical, mechanistic and/or observational/registry analyses and reviews suggesting plausible protective effects (i.e., improved endothelial function, reduced inflammation, improved metabolic milieu) in other populations.^29^ For OSA patients on PAP, GLP-1RA therapy appears complementary as PAP remains the gold standard for immediate control of obstructive events, while GLP-1RAs addresses the root cause (i.e., obesity) and can reduce AHI and hypoxic burden, sometimes allowing re-evaluation of pressure needs or even meaningful disease improvement. The SURMOUNT-OSA trials included a PAP-using subgroup with similar benefit. Nonetheless, long-term PAP adherence and real-world persistence are important limitations.^15^

### Observed superiority of tirzepatide versus semaglutide

The observed superiority of tirzepatide likely arises from its dual agonism of both GLP-1 and glucose-dependent insulinotropic peptide (GIP) receptors versus selective GLP-1RA agonism by semaglutide. Pooled analyses demonstrate larger mean weight loss with tirzepatide than semaglutide at 6 months, and a higher proportion of patients reaching ≥10% weight reduction with tirzepatide.^30^ This mechanism is associated with greater reductions in body weight, improved insulin sensitivity and more pronounced anti-inflammatory effects.

^31,32^ Such actions are particularly relevant to processes underlying pulmonary vascular remodeling and hypoxia-induced cardiopulmonary stress in individuals with obesity and OSA. GIP exerts direct vascular actions through functional GIP receptors expressed on endothelial cells, where ligand binding activates G-protein–coupled signaling cascades leading to increased intracellular cyclic AMP and protein kinase A (PKA) activation. The cAMP-PKA signaling framework engages downstream kinase pathways known to regulate endothelial homeostasis.^33^ Within the pulmonary circulation, such signaling intersects mechanistically with nitric oxide (NO), where endothelial nitric oxide synthase (eNOS) produces constitutive, low-level NO that maintains vascular tone, inhibits smooth muscle proliferation and preserves endothelial function.^34^ Activation of cAMP-PKA pathways is known to enhance eNOS activity through phosphorylation and improved coupling, thereby increasing NO bioavailability and cyclic GMP signaling in adjacent pulmonary vascular smooth muscle cells. In obesity, OSA and related cardiometabolic states, endothelial dysfunction characterized by impaired eNOS signaling, oxidative stress, and reduced NO bioavailability contributes to pulmonary vasoconstriction, vascular remodeling, and elevated pulmonary arterial pressures. Augmentation of endothelial incretin signaling, particularly through GIP receptor activation, may therefore restore NO-dependent vasodilatory capacity, attenuate hypoxia-induced pulmonary vasoconstriction, and limit maladaptive pulmonary vascular remodeling. These mechanistic links provide plausible pathways through which dual GIP/GLP-1 receptor agonists exert salutary effects on pulmonary vascular physiology beyond weight loss alone, potentially modifying the development and/or progression of pulmonary hypertension in high-risk populations.

#### Study strengths

To our knowledge, it represents the largest real-world comparative effectiveness analysis evaluating PAP therapy alone versus PAP combined with GLP-1 RAs, for incident pulmonary hypertension and all-cause mortality. Additional strengths include the use of a large, diverse population and rigorous analytic methods, including propensity score matching, which enhance internal validity while supporting generalizability to routine clinical practice.

#### Study limitations

First, analysis was limited to two GLP-1 RAs and did not evaluate dose-specific effects. Second, reliance on administrative diagnostic and medication codes may result in misclassification, particularly for PH and OSA. However, such misclassification is likely to be nondifferential across treatment groups and would be expected to bias results toward the null. Third, information on medication adherence, PAP compliance, and treatment-related adverse effects was not available and could have influenced outcomes. Fourth, although PSM was used to mitigate confounding, unmeasured confounding remains possible. Finally, restriction of outcome analyses to incident cases resulted in some exclusions after matching, an inherent TriNetX platform limitation in which prevalent case exclusion occurs after matching rather than before. Despite this limitation, the large analytic sample sizes retained for outcome analyses support the robustness of the findings.

## CONCLUSIONS

Obese-OSA patients treated with PAP taking GLP-1RAs exhibited significantly lower 1-year and 3-year incident PH and all-cause mortality versus PAP. Tirzepatide exhibited further lowering incident PH and mortality versus semaglutide, showing increased and sustained benefits over time.

## Data Availability

Data publicly available online at TriNetX Database

https://live.trinetx.com/

## Acknowledgments

The first author was responsible for data access, integrity of the data, and data analysis.

